# Verbal fluency tests assess global cognitive status but have limited diagnostic differentiation: Evidence from a large-scale examination of six neurodegenerative diseases

**DOI:** 10.1101/2022.08.16.22278837

**Authors:** Shalom K. Henderson, Katie A. Peterson, Karalyn Patterson, Matthew A. Lambon Ralph, James B. Rowe

## Abstract

**Objective:** Verbal fluency is clinically widely used but its utility in differentiating between neurodegenerative dementias and progressive aphasias, and from healthy controls, remains unclear. We assessed whether the total number of words produced, their psycholinguistic properties, and production order effects could differentiate between Alzheimer’s disease (AD), behavioural variant of frontotemporal dementia (bvFTD), non-fluent and semantic variants of primary progressive aphasia (PPA), progressive supranuclear palsy (PSP), corticobasal syndrome (CBS), and healthy controls.

**Methods:** Category and letter fluency tasks were administered to 33 controls and 139 patients at their baseline clinical visit: 18 AD, 16 bvFTD, 26 nfvPPA, 26 svPPA, 36 PSP, and 17 CBS. We assessed group differences for total words, psycholinguistic word properties, and associations between production order and exemplar psycholinguistic properties. Receiver Operating Characteristic (ROC) curves determined which measure could best discriminate patient groups and controls.

**Results:** Total word count distinguished controls from all patient groups, but neither this measure nor the word properties differentiated the patient groups. ROC curves revealed that, when comparing controls to patients, the strongest discriminators were total word count followed by word frequency. Word frequency was the strongest discriminator for svPPA versus other groups. Fluency word counts were associated with global severity as measured by Addenbrooke’s Cognitive Examination-Revised (ACE-R).

**Conclusions:** Verbal fluency is an efficient test for assessing global brain-cognitive health but has limited utility in differentiating between cognitively- and anatomically-disparate patient groups. This outcome is consistent with the fact that verbal fluency requires many different aspects of higher cognition and language.

## Introduction

Beyond brief clinician-rated global assessment instruments such as the Clinical Dementia Rating (CDR) scale, verbal fluency tests are one of the most widely used assessments in clinical and research settings. They are quick and easy to administer and score, require no assessment equipment, can differentiate between healthy populations and those with neurodegenerative disease, and are sensitive to cognitive or language decline [1, 2]. Verbal fluency tests assess an individual’s ability to generate words from a specified letter of the alphabet (e.g., F, A, S) or a semantic category (e.g., animals, fruits). Difficulty or errors may arise from impairments in one or more aspects of cognition including attention, working memory, semantic memory, executive functioning, and language. Verbal fluency deficits have been identified in Alzheimer’s disease (AD), frontotemporal dementia (FTD), vascular dementia, dementia with Lewy bodies, progressive supranuclear palsy (PSP), and other parkinsonian disorders [3-5]. Typically, previous studies have focussed on one diagnostic group to explore the qualitative and quantitative changes (e.g., total number of words, amount of clustering and switching) in the patients’ performance and pattern of words produced. Whereas the total word count may be an indicator of cognitive impairment, it remains unclear how well verbal fluency tests can differentiate all patient types from heathy controls or between patient groups [5, 6]. The current study used a large-scale transdiagnostic approach to examine letter and category fluency performance assessed at first clinical visit in six different clinical groups representing various cortical and subcortical neurodegenerative disorders including primary progressive aphasias (PPA). We asked some simple but clinically-important questions: (a) how well does verbal fluency performance distinguish each group from healthy controls at this first clinical visit; (b) do these tests contribute to differential diagnosis between patient groups; and (c) can differential diagnosis be improved if we move beyond total word count to more detailed analysis of the characteristics of the words produced?

The lexical-semantic features of words produced in tests of verbal fluency (e.g., frequency, imageability, age of acquisition) are less studied than other measures [7]. In semantic dementia (SD)/semantic variant of primary progressive aphasia (svPPA), connected speech production and naming show a relative over use of words that are more frequent, more abstract (i.e., “lighter” nouns and verbs) and earlier acquired [8-10]. This pattern is reported to a lesser extent in AD [11-13]. Although verbal fluency is severely reduced in PSP and corticobasal syndrome (CBS), the psycholinguistic properties of the words have not been investigated in detail: it has been proposed that PSP leads to production of few words of relatively low frequency [14-16]. With the exception of reduced word length observed in non-fluent variant PPA (nfvPPA) [17], it is unclear which word properties, if any, might be informative in classifying patients with nfvPPA, or other forms of FTD, compared to other disorders.

In healthy adults, production order, sometimes referred to as serial recall order, reveals a pattern of increasing lexical and semantic richness and difficulty [18, 19]; but whether this link applies in dementia and aphasia is unknown. Distinct patterns across diagnostic groups could potentially aid diagnostic differentiation and elucidate neurocognitive systems underlying verbal fluency.

This study tested the hypothesis that, in standard clinical versions of the verbal fluency task, the number of words, their psycholinguistic properties, and/or production order effects would differentiate neurodegenerative dementias and aphasias. We explored these features in direct comparisons across a large dataset collected from a broad range of patient groups including amnestic presentation of AD, behavioural and language variants of FTD, and the ‘parkinson-plus’ disorders of PSP and CBS.

The study had three specific aims: 1) to assess whether the total number of words produced during a verbal fluency task can differentiate between diagnostic groups, having controlled for individual differences in age, gender and education; 2) to identify the multivariate lexico-semantic features of words generated by patients, using a principal component analysis to reduce dimensionality; and 3) to determine the association between the item-level lexico-semantic features and their production order, for each diagnostic group.

## Materials and methods

### Participants and data acquisition

We analysed data from people with clinical diagnoses of AD (n=18) [20], behavioural variant of FTD (bvFTD; n=16) [21], CBS (n=17) [22], PSP (n=36) [23], nfvPPA (n=26) and svPPA (n=26) [24, 25], as well as healthy controls (n=33). Patients were recruited from specialist memory and movement disorders clinics at Cambridge University Hospitals NHS Trust in observational studies (REC references 07/Q0102/3; 10/H0308/34; 12/EE/0475; 14/LO/2045; 16/LO/1735). Demographic information including ACE-R and MMSE scores are presented in Table 2.

Verbal fluency tests were administered during the baseline visit. Participants were asked to name as many words as they could that (a) began with the letter ‘P’ (excluding people and place names) and (b) that belonged to the category of ‘animals’, to assess phonemic/letter and semantic/category fluency respectively. Words (including bigrams) were recorded over 60 seconds for each task and transcribed by the examiner. The total word count, excluding errors and repetitions, was calculated. For each of the words, we obtained ratings for psycholinguistic properties from the MRC Psycholinguistic Database [26] and the English Lexicon Project [27] as listed in Table 1. Where ratings for the pluralised word were unavailable, the word properties for the singular version were extracted.

**Table 1.** Word properties and definitions

| **Word Property** | **Definition** |
| --- | --- |
| Frequency | How many times a word appears in a corpus |
| Imageability | How well a word gives rise to a sensory experience or mental image (e.g., ‘pots’ is more imageable than ‘possibility’) |
| Age of Acquisition | When individuals typically learn a word in spoken or written form (e.g., ‘pterodactyl’ is acquired at a later age than ‘people’) |
| Familiarity | The degree to which an individual comes in contact with or thinks about the concept |
| Concreteness | The degree to which a word is considered more concrete or more abstract (e.g., the concept of ‘elephants’ is considered highly concrete whereas ‘paradox’ is abstract) |
| Length | Number of letters in a word |
| Orthographic Levenshtein Distance (OLD) | The number of insertions, deletions and substitutions needed to generate one letter string from another; the mean orthographic distance to the 20 closest orthographic neighbours (e.g., a word with low orthographic neighbourhood like ‘veil’ will have a higher OLD value based on the number of changes needed to create words like ‘boil’, ‘cell’, and ‘fell’ than a word with high orthographic neighbourhood like ‘lime’ to create words like ‘lie’, ‘dime’ and ‘limb’) |
| Phonological Levenshtein Distance (PLD) | The mean number of steps required through phonemic substitutions, insertions, or deletions to transform a word into its 20 closest phonological neighbours (e.g., a word with a distant phonological neighbourhood like ‘insomnia’ will have a high PLD based on the number of changes needed to create words like ‘insignia,’ ‘inertia,’ and ‘anaemia’ relative to a word like ‘resume’ to create words like ‘result,’ ‘refute,’ and ‘legume’) |
| Semantic Neighbourhood Density (SND) | The proximity of semantic neighbours to a target word (e.g., an animal such as ‘dog’ has a low semantic distance with a dense neighbourhood since its nearest neighbours such as a ‘cat’ would be relatively close to the target word, as compared to an animal like a ‘kangaroo’) |
| Semantic Diversity | The degree to which the different contexts associated with a given word vary in their meanings (e.g., words such as ‘place’ or ‘people’ have higher values as they appear in more diverse context than words like ‘peacock’) |

### Statistical analysis

Between-group differences for total word count were tested by two-way analysis of covariance (ANCOVA) with age and sex as covariates. *Post hoc* analyses were conducted using Tukey HSD for multiple comparisons. To establish whether verbal fluency can indicate not only the presence, but also the severity, of global cognitive impairment, we computed correlations across participants to assess two forms of associations: (1) between ACE-R and total word count, plus word counts for letter and category fluency separately, and (2) between letter and category fluency.

Next, average ratings per participant for the ten word properties listed in Table 1 were entered into a principal component analysis (PCA). A Kaiser-Meyer-Olkin test determined the suitability of our dataset for PCA. We selected three components based on Cattell’s criteria and then performed varimax rotation. Using factor scores per participant, we conducted a one-way analysis of variance (ANOVA) to test for group differences followed by *post hoc* analyses using Tukey HSD for multiple comparisons.

For item-level ‘production order-psycholinguistic feature’ scoring, each word was scored according to its production order position and the three psycholinguistic features that were individually most strongly associated with principal components 1 to 3 – namely: length, imageability and word frequency. For each diagnostic group and over the whole study population, correlation coefficients (Spearman’s rho) were calculated between the production order and these three psycholinguistic features (Figure 4). To capture individual variability of strength and direction, linear regression analyses were run to assess the relationship between the production order and the aforementioned features for each participant. Beta coefficients were extracted to test within and between group differences via a six group x two fluency type ANOVA.

Lastly, logistic regression analyses were conducted to ascertain which measure could best discriminate different patient groups and controls. Receiver Operating Characteristic (ROC) curves were generated using the pROC package [28]. All statistical analyses were performed in R statistical software.

## Results

### Demographics

Demographic details are shown in Table 2. *Post hoc* tests confirmed that, as expected, patients with bvFTD were younger than those with CBS (*p* < 0.001), nfvPPA (*p* < 0.001), PSP (*p* < 0.001), and controls (*p* = 0.02). No other groups differed in age. There were no significant differences between groups in terms of baseline education, handedness or gender. There were significant differences between groups on the ACE-R and MMSE, with controls performing better than all patient groups on both tests. Patients with PSP scored higher on the ACE-R than those with svPPA (*p* = 0.02) and AD (*p* = 0.02). Patients with AD scored lower on the MMSE than those with svPPA (*p* = 0.01), nfvPPA (*p* = 0.05), and PSP (*p* = 0.004).

**Table 2.**
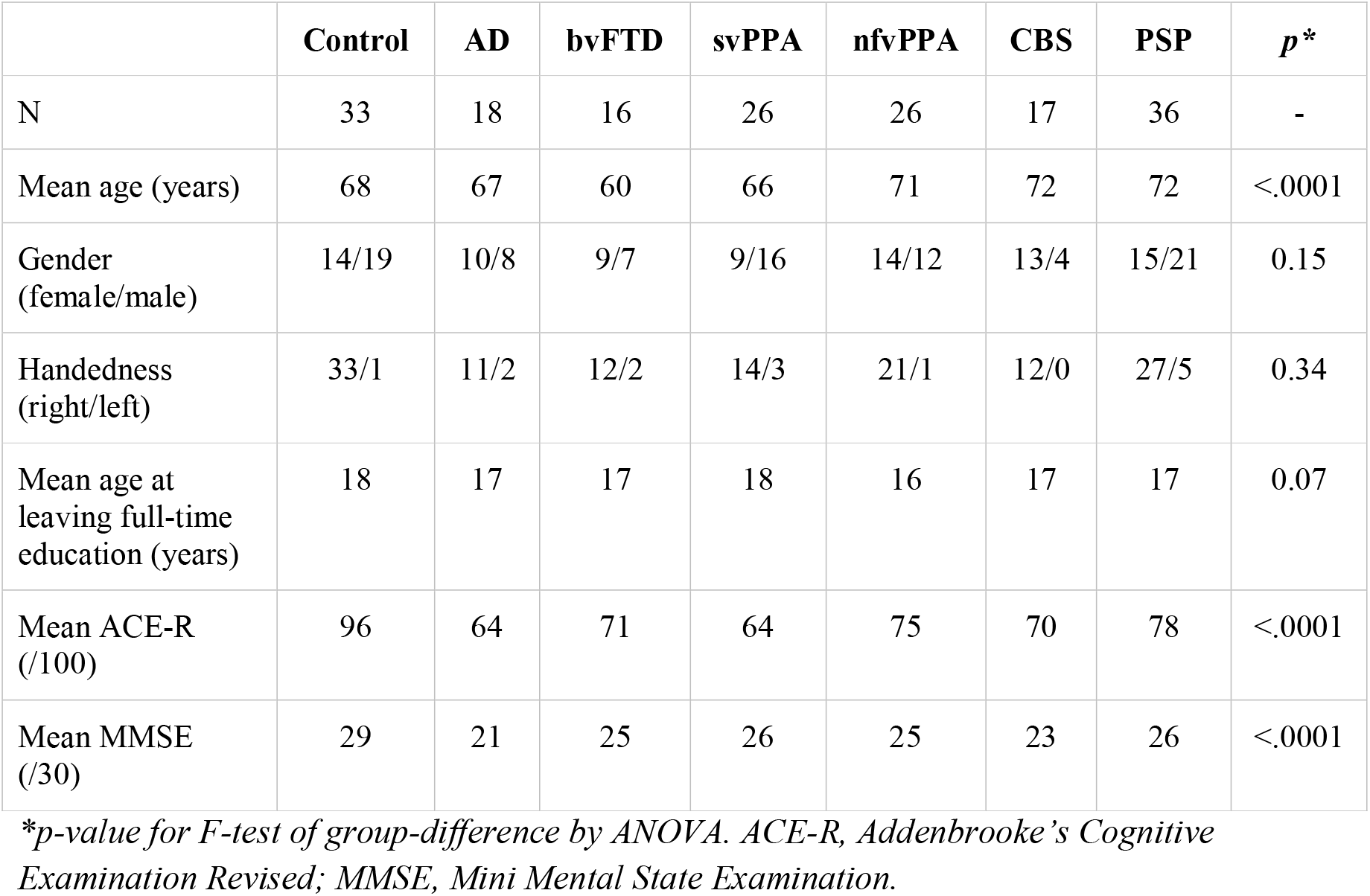
Demographics for the study population recorded at the baseline visit

### Diagnostic differentiation

#### 1. Total word count

As shown in Figure 1, there was a significant effect of group on the total word count, in both letter and category tasks, after controlling for age and sex. Specifically, there was a significant effect of group (F(6,324) = 68.3, *p* < .001), fluency type (F(1,324) = 33.5, *p* < .001), and group-by-fluency type interaction (F(6,324) = 2.4, *p* = .03), indicating that (a) controls produced more words than all patient groups (control vs. each patient group, *p* < .001) and (b) more words were produced during category relative to letter fluency. *Post hoc* analyses with Tukey HSD multiple comparisons correction revealed a significant difference between the total word count in letter versus category fluency in controls (*p* < .001) and (marginally significant) in PSP (*p* = 0.06).

**Figure 1.**
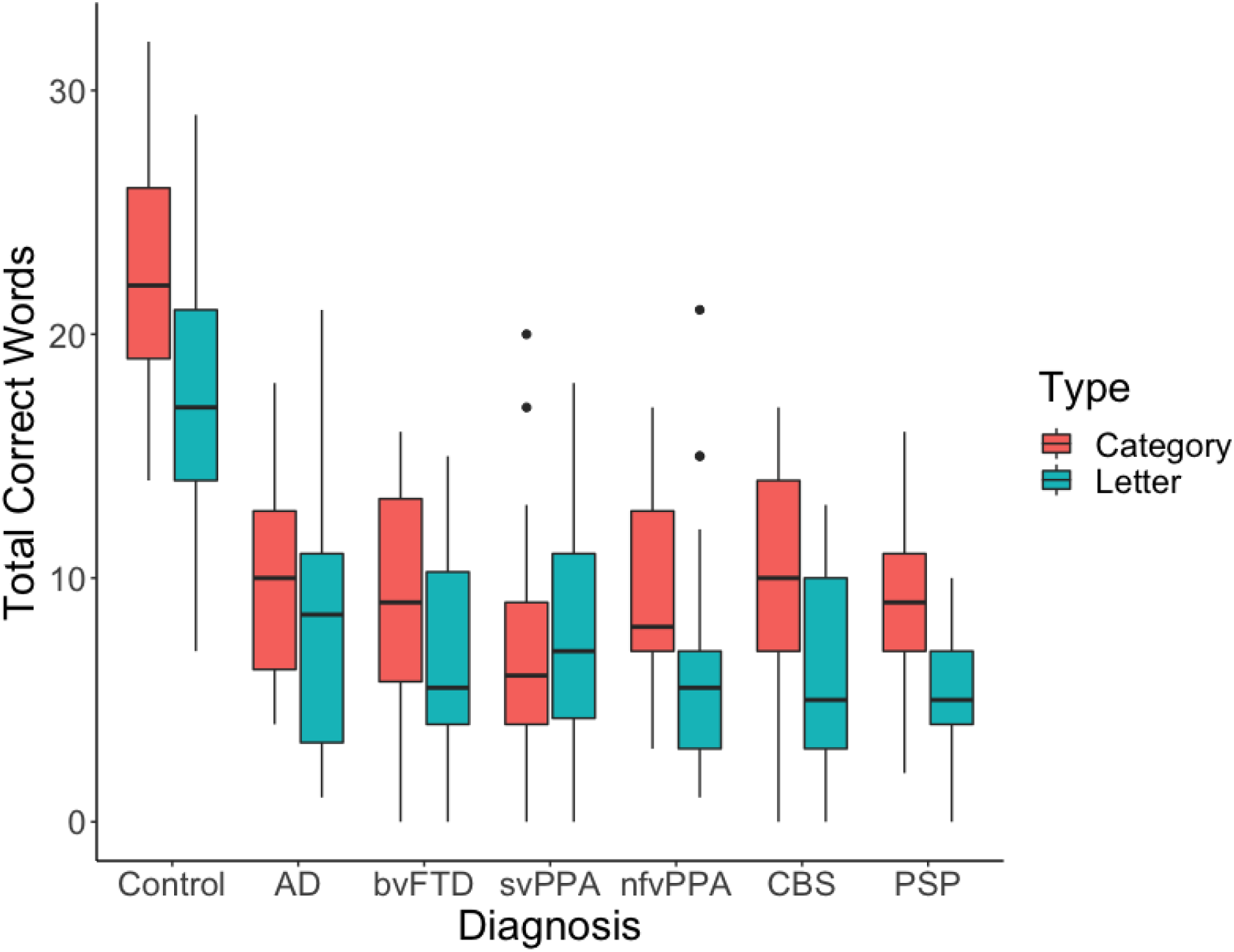
Total words produced by controls and patient groups during category and letter fluency. Group data illustrate healthy controls performinig better than all patient groups.

#### 2. Associations between global severity, letter, and category fluency

A pearson correlation coefficient was computed to assess the linear relationship between (i) ACE-R as a measure of global severity, and total word count, word count for letter and category fluency, and (ii) word count for letter versus category fluency. As shown in Figure 2, ACE-R was positively correlated with total word count, r(138) = 0.49, *p* < 0.001, letter fluency word count, r(138) = 0.43, *p* < 0.001, and category fluency word count, r(138) = 0.50, *p* < 0.001. A positive correlation was also found between letter and category fluency, r(138) = 0.79, *p* < 0.001.

**Figure 2.**
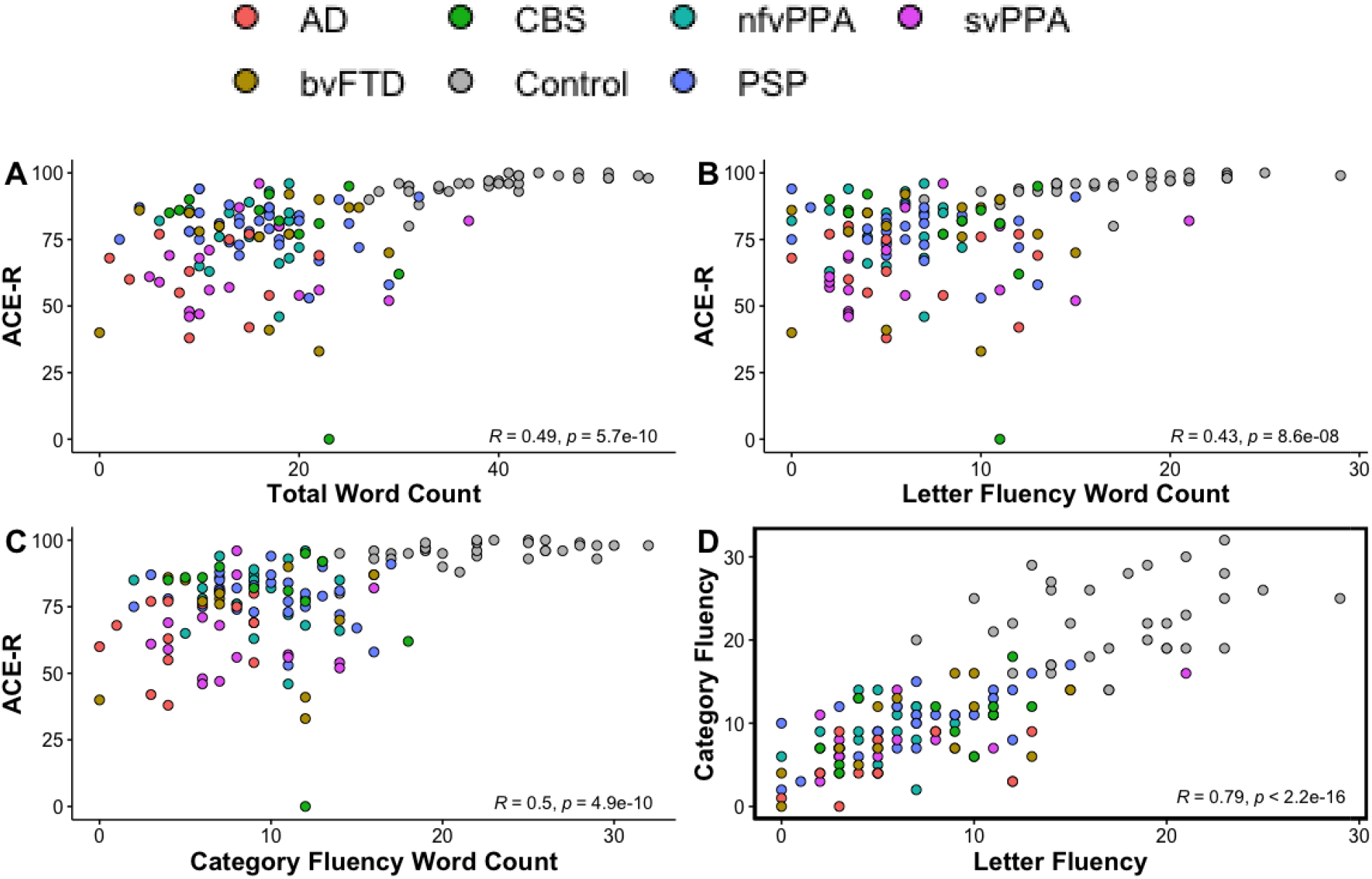
Associations between ACE-R and word counts and between letter and category fluency. (A) Correlation between ACE-R and total word count; (B) Correlation between ACE-R and word count for letter fluency; (C) Correlation between ACE-R and word count for category fluency; (D) Correlation between words counts for letter versus category fluency.

#### 3. Syndromic dimensions of word properties using principal components analysis

Mean ratings for each word property per participant, excluding controls, were entered into a PCA with varimax rotation. Three principal components were identified using Cattell’s criteria, each representing a group of covarying psycholinguistic features of words produced by patients. Three components explained 87.3% of the variance (Kaiser-Meyer-Olkin = 0.77). The loading of each measure is given in Table 3.

**Table 3.** Loadings for PCA of word properties

| Measure | PC 1 (Phonological Length) | PC 2 (Semantic Richness) | PC 3 (Lexical Familiarity) |
| --- | --- | --- | --- |
| Length | **0.91** | -0.24 | -0.28 |
| OLD | **0.93** | 0.00 | -0.33 |
| PLD | **0.93** | 0.00 | -0.29 |
| Concreteness | 0.00 | **0.95** | 0.00 |
| Imageability | 0.00 | **0.96** | 0.00 |
| Age of acquisition | 0.38 | **-0.66** | **-0.55** |
| Semantic diversity | 0.13 | **-0.60** | **0.50** |
| Frequency | **-0.50** | 0.00 | **0.78** |
| Familiarity | -0.36 | 0.20 | **0.78** |
| SND | -0.49 | -0.13 | **0.77** |

*Rotation: Orthogonal varimax. Loadings above a threshold of 0.5 are bolded. PC, principal component; OLD, orthographic levenshtein distance; PLD, phonological levenshtein distance; SND, semantic neighbourhood density.*

Principal component (PC) 1 (see Figure 3A) was loaded heavily by length, OLD and PLD, and was thus interpreted as ‘phonological length’ where positive scores reflect a greater production of words that are longer (e.g., ‘prescription’ and ‘psychological’ vs ‘pan’ and ‘pen’) with high phonological and orthographic levenshtein distance (e.g., ‘plum’, ‘premature’, and ‘proliferate’ vs ‘pen’, ‘pan’ and ‘pat’). The results from a one-way ANOVA revealed significant group differences in PC 1 (F(5,131) = 3.38, *p* = 0.007), driven by patients with svPPA producing shorter and phonologically less complex words than patients with PSP (*p* = 0.008) and CBS (*p* = 0.02).

**Figure 3.**
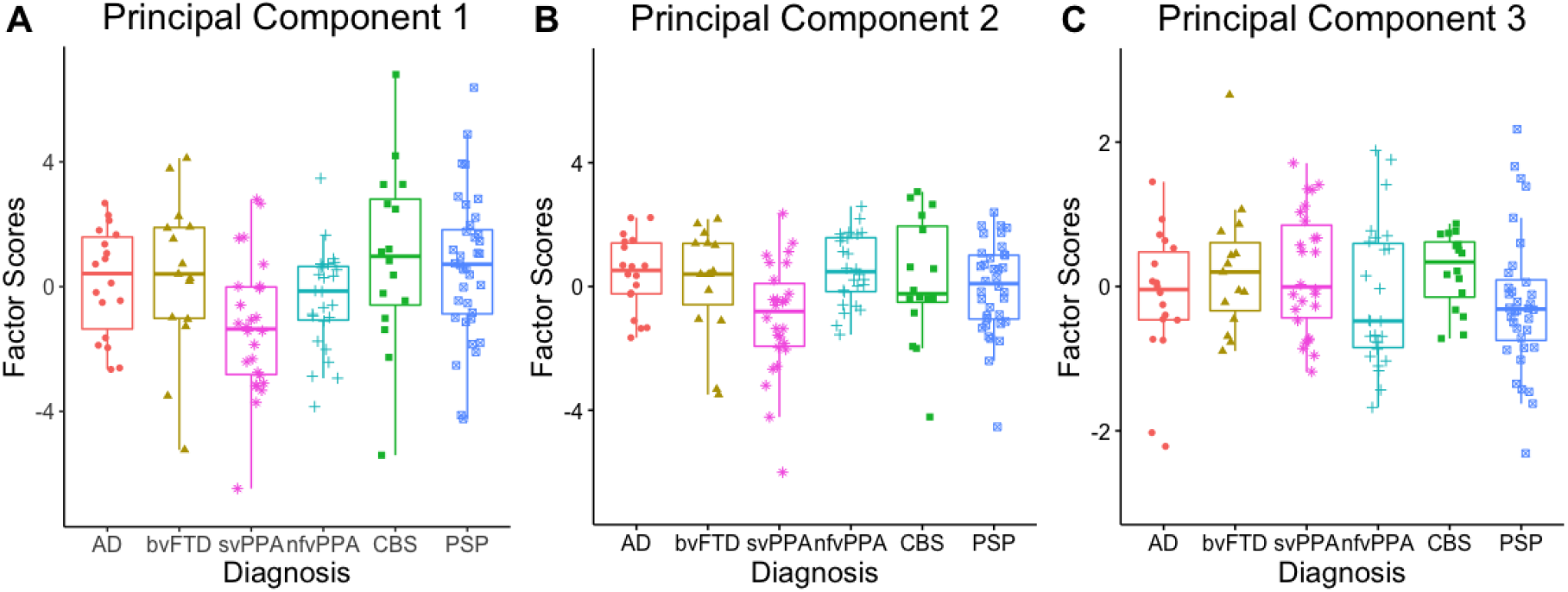
Principal component analysis scores of word properties across diagnostic groups. (A) PC 1: ‘phonological length’; (B) PC 2: ‘semantic richness’; (C) PC 3: ‘lexical familiarity’. AD, Alzheimer’s disease; bvFTD, behavioural variant frontotemporal dementia; svPPA, semantic variant primary progressive aphasia; nfvPPA, non-fluent variant primary progressive aphasia; CBS, corticobasal syndrome; PSP, progressive supranuclear palsy.

PC 2 (Figure 3B) was interpreted as ‘semantic richness’ since concreteness, imageability, age of acquisition, and semantic diversity loaded heavily on it. Positive scores represented a greater production of highly imageable and concrete words that were acquired earlier and have less semantically diverse meanings (e.g., ‘cow’ and ‘elephant’ vs. ‘fowl’ and ‘louse’). Significant group differences were found for PC 2 (F(5,131) = 3.20, *p* = 0.009), driven by patients with svPPA producing less concrete, less imageable, and later acquired words relative to patients with nfvPPA (*p* = 0.005) and AD (*p* = 0.04).

PC 3 (Figure 3C) was loaded heavily by lexical-semantic features including frequency, familiarity, and semantic neighbourhood density, and was interpreted as ‘lexical familiarity’. Positive scores represented a greater production of more frequent and familiar words with higher semantic density (e.g., ‘dog’ and ‘fish’ vs. ‘tiger’ and ‘snake’). A one-way ANOVA did not reveal significant group differences for PC 3 (F(5,131) = 0.300 *p* = 0.42).

#### 4. Item-level fluency with production order

Correlation coefficients (Spearman’s rho) were calculated for each participant between the production order (PO) and the three psycholinguistic features that were individually most strongly associated with principal components 1 to 3, namely length, imageability and frequency. Figure 4 plots the ‘PO-psycholinguistic feature’ trends averaged across groups. The ‘PO-length’ effect was significantly positive for letter (*p* = 0.002) and category (*p* < 0.001) over the whole study population, and for each group except letter fluency in CBS (r = -0.06); although CBS had the most positive correlation for category fluency (r = 0.46). The ‘PO-imageability’ effect was significantly negative for letter (*p* < 0.001) and category (*p* < 0.001) over the whole study population and for each group. The most negative correlations were found in bvFTD for letter fluency (r = -0.39) and nfvPPA and svPPA for category fluency (r = -0.31). The ‘PO-frequency’ effect was significantly negative for both letter (*p* < 0.001) and category (*p* < 0.001) over the whole study population and for each group except letter fluency in CBS (r = 0.03) and bvFTD (r = 0.03). Patients with svPPA showed the most negative correlation during category fluency (r = -0.44).

**Figure 4.**
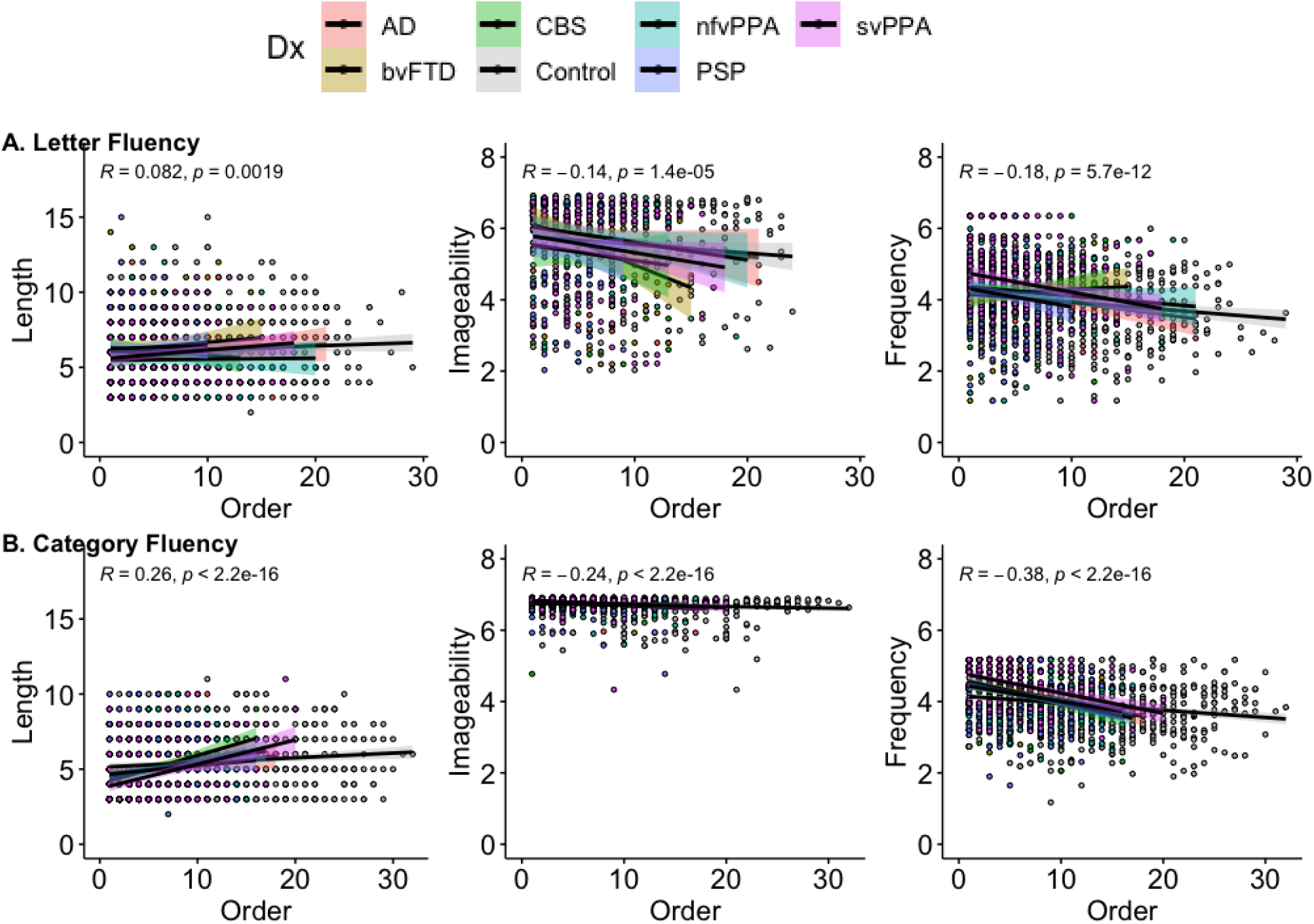
Production Order (PO)-word feature scatterplots over the whole study population. (A) PO-Length (left), PO-Imageability (middle), and PO-Frequency (right) for letter fluency; (B) PO-Length (left), PO-Imageability (middle), and PO-Frequency (right) for category fluency.

Linear regression analyses were used to assess the relationship between the PO and length, imageability, and frequency for each participant and beta coefficients were extracted to test within and between group differences. Six groups x two fluency type ANOVAs failed to reveal any significant effect of group or type for length (group: F(6,314) = 1.34, *p* = 0.24); type: F(1,314) = 2.28, *p* = 0.13), imageability (group: F(6,299) = 1.34, *p* = 0.24; type: F(1,299 = 1.53, *p* = 0.87) or frequency (group: F(6,315) = 1.72, *p* = 0.12; type (F(1,315) = 2.82, *p* = 0.09).

#### 5. Logistic regression

Logistic regression analyses were conducted to determine which measures (i.e., total word count and/or the psycholinguistic properties associated with these words) could discriminate (i) controls *versus* all patient groups, and (ii) between one patient group and the others. The discrimination between groups is reported as ROC curves in Figure 5.

**Figure 5.**
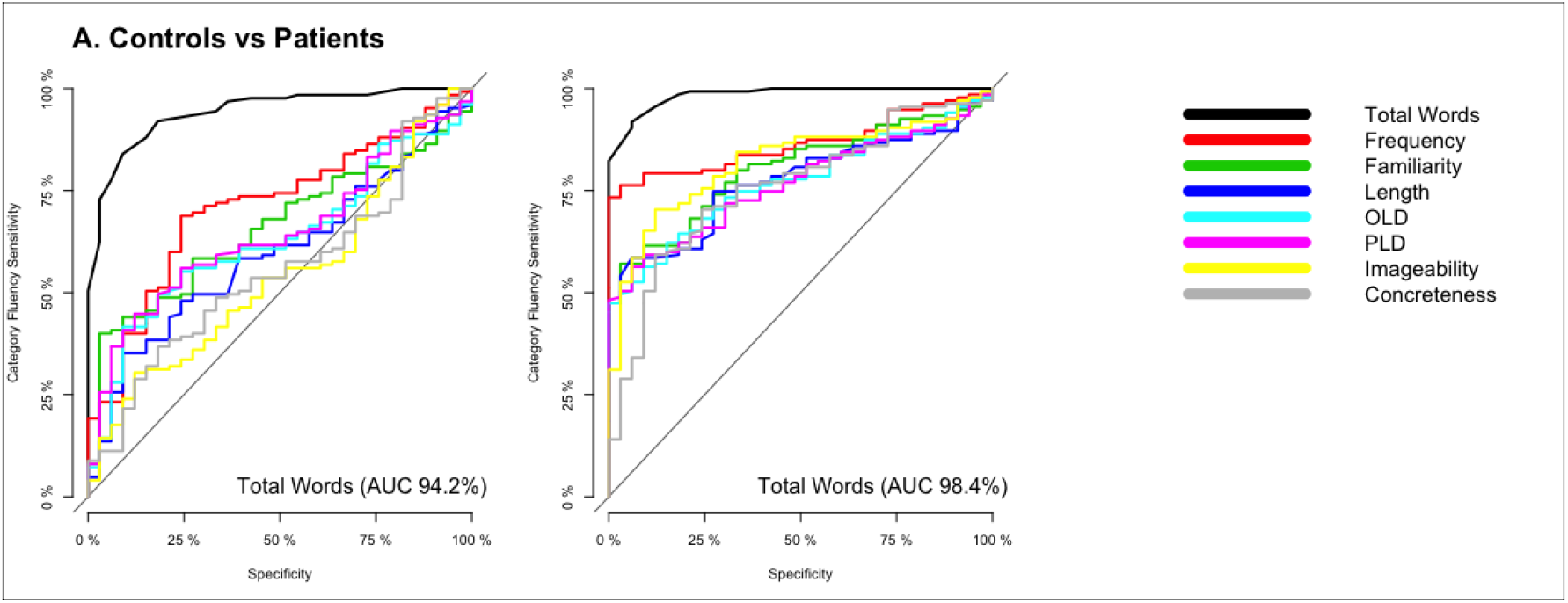
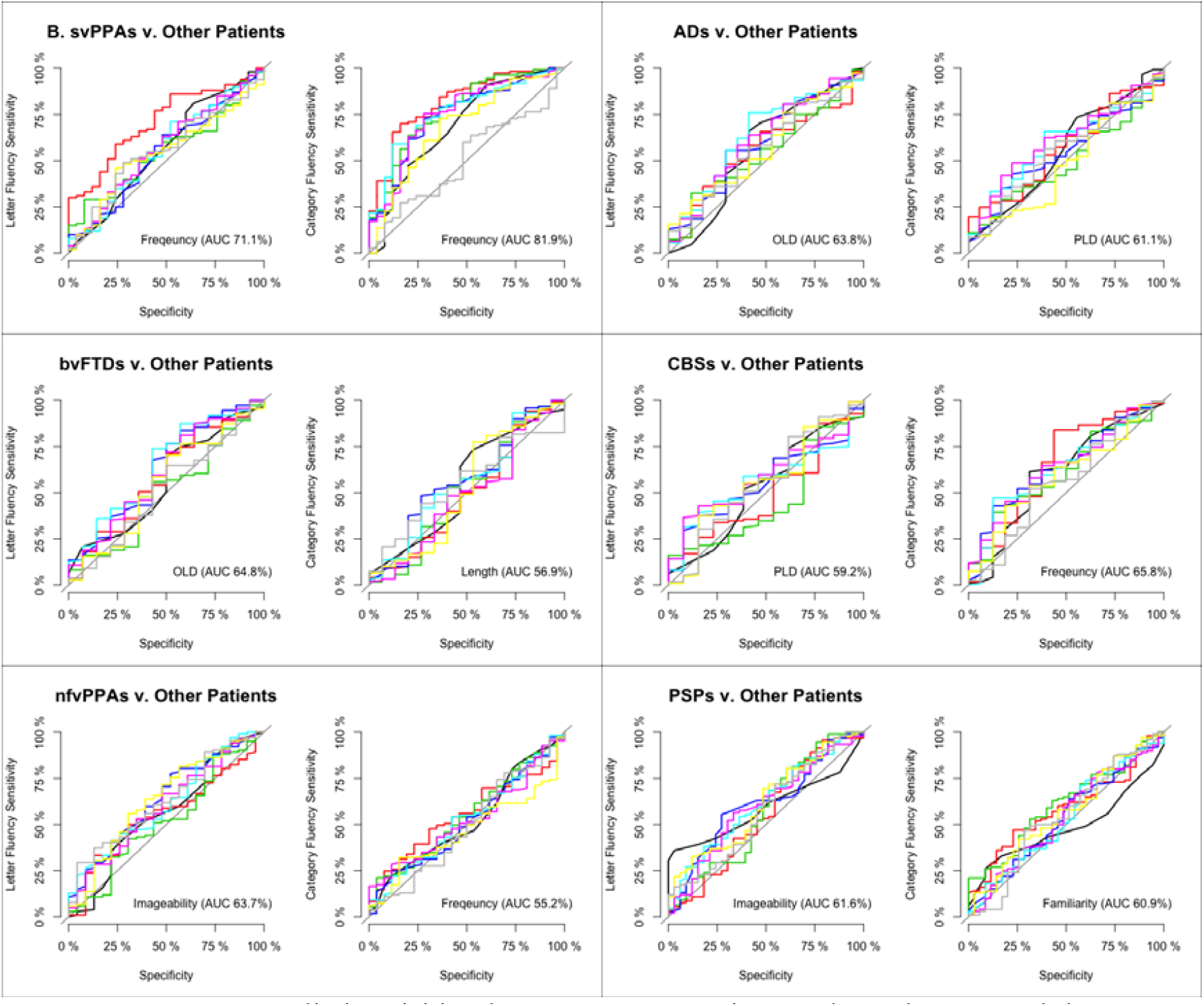
ROC curves distinguishing between groups using total word count and the psycholinguistic properties of these words during letter and category fluency. (A) Controls versus all patient groups; (B) Each patient group against other patient groups.

As shown in Figure 5A, when comparing controls relative to all patients, the strongest discriminator for letter fluency was total word count (AUC = 94.2%) followed by word frequency (AUC = 71.1%). For category fluency, the strongest discriminators were also total word count (AUC = 98.4%) and word frequency (AUC = 86.6%).

The fluency metrics did not discriminate between most patient groups with high accuracy (see Figure 5B). A partial exception was the moderate discrimination of svPPA against all other patient groups with word frequency as the strongest discriminator for letter (AUC = 71.1%) and category (AUC = 81.9%) fluency. The discrimination of each of the other patient groups was weak, at best. OLD (AUC = 63.8%) and PLD (AUC = 61.1%) were the best measures for AD patients in letter and category fluency. OLD (AUC = 64.8%) and length (AUC = 56.9%) were also the best measures for bvFTD patients in letter and category fluency. For CBS patients, PLD (AUC = 59.2%) and frequency (AUC = 65.8%) were the best discriminators for letter and category fluency. Imageability was the best variable for letter fluency for both nfvPPA and PSP patients (AUC > 61%); for category fluency, frequency (AUC = 55.2%) and familiarity (AUC = 60.9%) were best discriminators for patients with nfvPPA and PSP, respectively.

## Discussion

Although verbal fluency tests are one of the most popular assessments regularly administered in clinic, how well they can differentiate either all patient types from healthy controls or between patient groups has not previously been established. Furthermore, there has been little or no exploration in previous studies of whether differential diagnosis between various forms of cortical and subcortical neurodegenerative diseases can be improved with quantitative, albeit time-consuming, analyses of properties of the words produced. Using a large dataset collected from a broad range of neurodegenerative patient groups, we addressed these issues by examining letter and category fluency assessed at the first clinical visit using quantification of total word count and analysis of the qualities of the words produced. There were two very clear principal results. (1) The number of words produced in letter and/or category fluency strongly differentiated healthy controls from each neurodegenerative disease (amnestic AD, behavioural and language variants of FTD, and both PSP and CBS) and was associated with the severity of the patients’ global decline (as measured by ACE-R). (2) On the other hand, neither the total word count nor the psycholinguistic properties of the words produced differentiated between disorders, with the partial exception of svPPA (see below). These results are in line with previous studies of individual patient groups, which found that the total number of words produced can differentiate healthy controls from those with major neurocognitive disorders [29], as well as those with mild cognitive impairment from advanced dementias [30]. It seems very likely that this lack of diagnostic differentiation is because verbal fluency taxes multiple aspects of higher cognition and language. Thus, if any aspect of language, memory, attention or executive functioning is impaired, then performance on verbal fluency will be compromised regardless of specific diagnosis.

Going beyond the traditional measure of the number of words produced, we examined the psycholinguistic characteristics of the words produced by each patient group. These additional psycholinguistic measures showed weak differences between diagnostic groups (Figure 5). The only partial exception is that word frequency was a moderately strong discriminator in letter (AUC = 71.1%) and category (AUC = 81.9%) fluency for patients with svPPA. These cases were more likely than the other groups to generate items with higher word frequency, which aligns with the shift of word frequency observed in svPPA naming and connected speech [8-10]. Beyond this moderate effect, our results indicate that only subtle (non-significant), graded differences in lexico-semantic features are found at the group level and are unlikely to provide diagnostic differentiation for individual patients. From a clinical perspective it is also worth noting that examining the psycholinguistic properties of the words produced by each patient is laborious and would seem to have little clinical utility in light of the subtle differences between the diagnostic groups.

In addition to the total number of words produced, the type of verbal fluency has often been proposed to differentiate between different kinds of neurodegenerative disorder. Previous studies have reported either equally impaired performance on both types of fluency or better performance on category than letter fluency in patients with FTD, PSP, and CBS [15, 31, 32]. However, reports of unequal performance as a function of fluency type might merely reflect the normative pattern found in healthy controls [33]. A reverse of this pattern has been reported in patients with AD and SD [34-36], and thus it has been proposed that disorders that disrupt semantic memory will result in a more pronounced deficit for category relative to letter fluency [15]. This phenomenon is thought to arise from a reduction in the availability of semantic attributes following temporal lobe degeneration [37-40]. These past studies have typically focussed on an individual disorder, rather than the parallel systematic examination conducted in this study. Given that the multiple patient groups included in our study have characteristic differential anatomical distributions and associated variations in cognitive-language profiles, then we might have expected to observe contrastive effects of fluency type across the groups. Our results, however, only revealed significantly more words produced in category than letter fluency in healthy controls and (marginally) in patients with PSP. Indeed, the total numbers of words produced in each type of fluency were highly correlated (Figure 2D) indicating that they primarily rely on the same cognitive and language processes and to very similar degrees. Thus, our findings question the accuracy and clinical utility of discrepancy between category and letter fluency for diagnostic differentiation. There were some limitations to our study. First, we only present clinical, not pathological, diagnoses. Future studies of performance on fluency tasks might explore whether performance relates in any way to the type of pathology as well as the clinical diagnosis. Secondly, we did not directly explore the atrophy correlates of fluency performance. Future work could investigate the relationship between fluency performance and the level/distribution of atrophy in each patient group and in the clinical population as a whole. On the other hand, perhaps the more striking observation is that – despite considerable variations in the types of neurodegenerative patient groups included in our study – there was so little evidence of substantial variations the number or the pattern of words elicited. This global result suggests that, instead of fluency performance having clear and restricted atrophy correlates, multiple brain systems including cortical and subcortical regions are engaged by tests of verbal fluency. Although prior lesion and functional imaging studies have proposed that distinct brain areas support verbal fluency (e.g., prefrontal executive regions), even brief cognitive deconstruction of the fluency task implicates numerous language, memory and executive systems in good performance. As a result, it is perhaps unsurprising that the key result from this study is that verbal fluency is an excellent, efficient clinical task for assessing the presence and level of global brain-cognitive decline (i.e., differentiates patients from controls and more from less advanced disease) but is very limited in its utility to differentiate between cognitively- and anatomically-disparate patient groups.

## Conclusion

Our results have important clinical and research implications. The study supports previous claims that verbal fluency tests are clinically efficient and sensitive for detecting cognitive changes across many different types of neurodegenerative condition. In contrast, there was very limited evidence that fluency performance (e.g., total word count) can assist differential diagnosis. Indeed, even detailed investigation of word properties and order of production did not improve diagnostic differentiation; and such analyses are time consuming and impractical in clinical settings.

## Data Availability

All data produced in the present study are available upon reasonable request to the authors.

## Acknowledgements

We thank our patients and their families for supporting this work. For the purpose of open access, the author has applied a CC BY public copyright licence to any Author Accepted Manuscript version arising from this submission. The views expressed are those of the authors and not necessarily those of the NHS, the NIHR or the Department of Health and Social Care.

## Statements and Declarations

### Ethical standards statement

This study has been approved by the appropriate ethics committee and has been performed in accordance with the ethical standards laid down in the 1964 Declaration of Helsinki and its later amendments.

